# Enhancing Recruitment Rate and Heterogeneity in the TrialNet Pathway to Prevention Study: Insights from a Single Center Experience

**DOI:** 10.1101/2025.05.08.25327263

**Authors:** Alejandro F. Siller, Serife Uysal, Rebecca Schneider Aguirre, Tracy Patel, Ananta Addala, Laura M. Jacobsen, Heba M Ismail, Maria J. Redondo, Mustafa Tosur

## Abstract

Studies to predict and prevent type 1 diabetes (T1D) are limited by small numbers and lack of participation from all affected populations. We compared TrialNet Pathway to Prevention recruitments at one pediatric center 12 months before (June 2021-May 2022, n=15) and after (June 2022-May 2023, n=319) implementation of knowledge-based interventions aimed at clinicians and participants to increase recruitment. Recruitment increased by over 20-fold, with higher enrollment for all reported races and ethnicities although the overall population was largely non-Hispanic and White. Effective interventions are still needed to overcome recruitment barriers in order to ensure that the enrolled population reflects the overall affected population.

## Introduction

Improved understanding of the pathophysiology and natural progression of type 1 diabetes (T1D) has led to novel research into therapeutics designed to prevent T1D and its progression after onset. A recent notable milestone is the US Food and Drug Administration’s (FDA) approval of teplizumab, a monoclonal antibody treatment for delay of progression of T1D from stage 2 to stage 3 T1D [1]. Such advances have benefited from large multicenter trials recruiting both individuals with T1D and those identified as having risk factors for developing T1D in their lifetime. One example is Type 1 Diabetes TrialNet, an international consortium of physicians, scientists, and healthcare teams conducting research aimed at slowing or stopping the progression of T1D [2]. A major limitation of such studies, however, is that the generalizability of results depends on the heterogeneity of the enrolled population. In the case of diabetes, different phenotypes exist among individuals with a clinical diagnosis of T1D, including seronegative T1D [3] and T1D with inherited genetic markers of T2D that may represent overlap in phenotypes[4] to name just two examples. Such phenotypic variability poses challenges in the accurate diagnosis of diabetes type, especially in youth [5]. Variability in prevalence of confounding characteristics, such as obesity and insulin resistance, also exists among different racial/ethnic groups [5]. Robust recruitment of a heterogeneous population is therefore important in order to capture the full-range of disease manifestations, response to intervention, rates of side effects, and other important clinical characteristics. This paper reports on a single-center effort to boost TrialNet recruitment, and the demographics of the resulting cohort of enrollees.

## Methods

This is a retrospective study of registration data from the TrialNet Pathway to Prevention (PTP) study (NCT00097292). The PTP study recruits relatives of individuals with T1D who do not themselves have T1D and screens these relatives for islet autoantibodies, such as those against glutamic acid decarboxylase (GAD-65), IA2 antigen (IA2), insulin, and zinc transporter 8 (ZnT8). Participants in PTP can enroll through TrialNet’s online portal (www.trialnet.org/participate) or in person at a clinical site. Inclusion criteria during the time period from which data were obtained include being a first degree relative without diabetes aged between 2.5 and 45 years, or a second degree relative without diabetes aged between 2.5 and 20 years.

This study focused on registration data from Baylor College of Medicine/Texas Children’s Hospital, where strategies aimed at boosting recruitment were introduced in June 2022. **Figure 1** lists these strategies, sorted into which barrier to recruitment they targeted.

**Figure 1.**
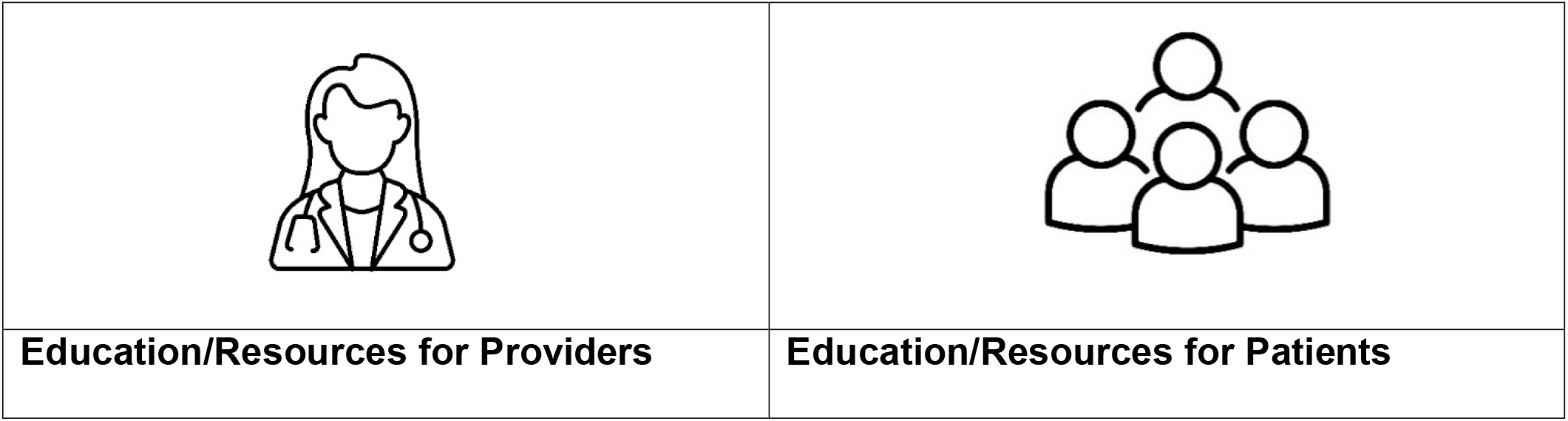
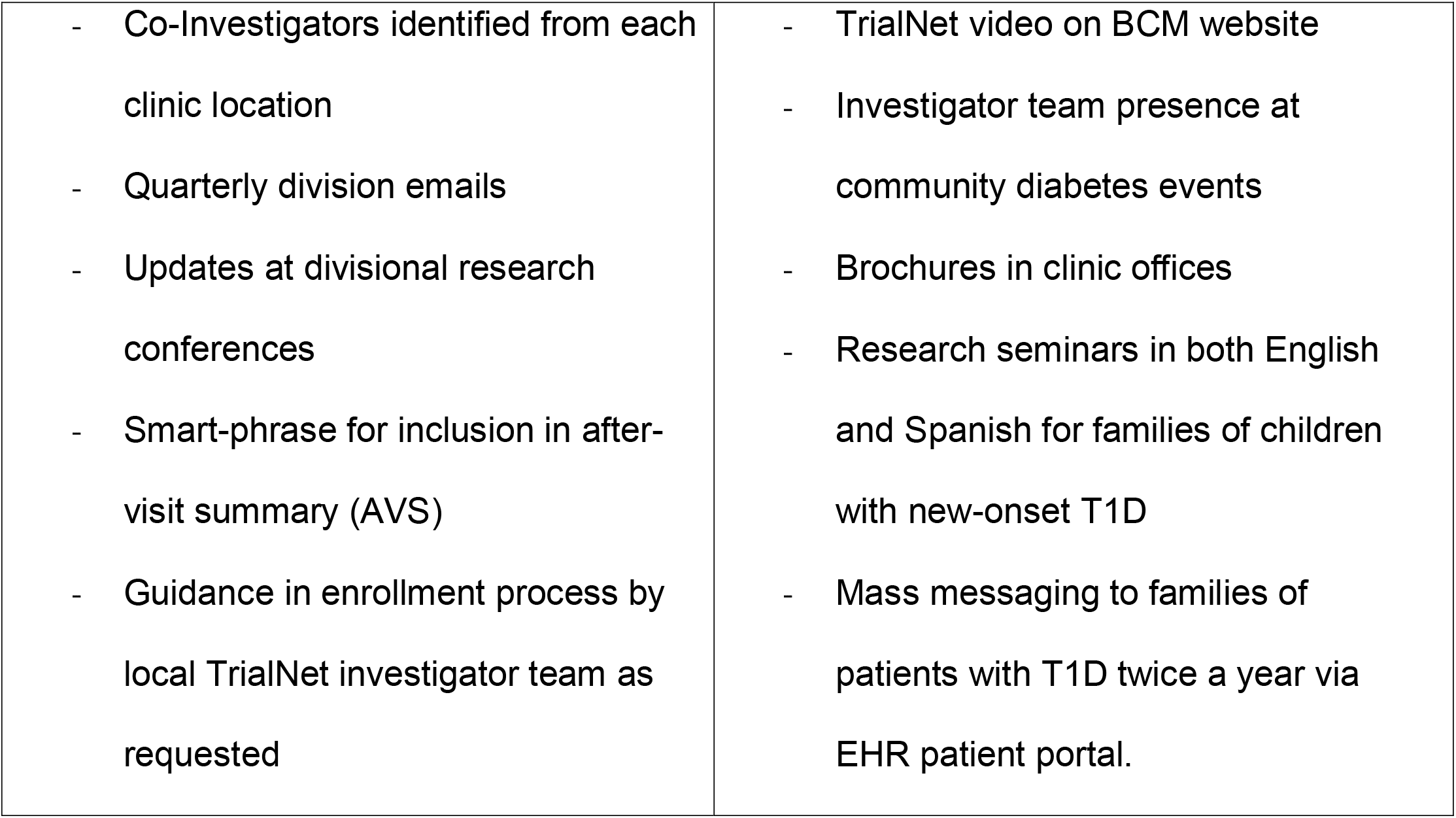
The multiple interventions adopted by one TrialNet site to boost recruitment into the Pathway to Prevention study. Interventions broadly aimed at a) educating providers about the benefits of participation in Pathway to Prevention, and how to enroll their patients’ eligible relatives, and b) educating families of patients directly on how to enroll eligible relatives. For providers, quarterly emails to our division provide a brief description of the services offered by TrialNet to participants and potential benefits, a list of providers who are also local co-Investigators in TrialNet at our various clinic sites who can be useful sources of information, and give running updates on progress in recruitment. EHR smart phrases allow providers less familiar with TrialNet or those who are short on time to add information to a patient’s after-visit summary about TrialNet including the TrialNet website where they can go to learn more. Patient-centered interventions included having literature available in the form of posters and brochures at every clinic, information seminars held for the families of newly-diagnosed children held in both English and Spanish, presence by our investigators at local community events hosted or attended by our division, inclusion of an informational video provided by TrialNet on the division’s web page, and direct messaging to patients via the EHR with information about TrialNet.

The investigators obtained registration data spanning from June 1, 2021 through May 31, 2023 from the TrialNet Data Coordinating Center. The selected dates encompassed registration numbers for 12 months before and after the implementation of these recruitment strategies. Data obtained from TrialNet were de-identified and included registration date, age at registration, sex, ethnicity, and race. Race and ethnicity were determined by self-report and using standard NIH classifications for race (American Indian or Alaska Native, Asian, Black or African American, White and More Than One Race) and ethnicity (Hispanic vs non-Hispanic) [6]. Data were limited to registrations in which the individual identified Baylor College of Medicine and Texas Children’s Hospital as the referring site.

We described the the race and ethnicity distributions of the pre- and post-intervention study cohorts, as well as that in the Texas Children’s Hospital Diabetes Center patient population, where recruitment was conducted. The small sample size of the pre-intervention cohort (n=15) precluded meaningful direct comparisons between the cohorts. Descriptive statistics of the cohorts were carried out using IBM SPSS Statistics for Mac, version 29.0.2.0 (IBM Corp., Armonk, N.Y., USA). All study participants provided informed consent prior to screening and enrollment, and the study was approved by a central Institutional Review Board (IRB), OneIRB at the University of South Florida, with IRB at each clinical site approving the reliance on the central IRB as the IRB of record.

## Results

Our site registered 334 individuals at risk for T1D during the study period (**Table 1**). The median (IQR) age was 13 (25.25) years and 53.2% were female. Racial distribution was 80.8% White, 3.6% Black, 2.7% Asian, 0.3% American Indian or Alaska Native and 12.6% more than one race or unknown, and 19.8% were Hispanic.

**Table 1.** Demographic characteristics of the overall population of registrants to TrialNet Pathway to Prevention study from our site during the study period, with breakdown of characteristics for the two periods, i.e., pre-intervention (from June 1, 2021 through May 31, 2022), and post-intervention (from June 1, 2022 through May 31, 2023).

|  | June 2021-May<br>2022 | June 2022-May<br>2023 | Overall Clinic<br>Population |
| --- | --- | --- | --- |
| Total Registrations | 15 | 319 | N/A<br>(total patient<br>population 3598) |
| No. Female Registrants (%) | 7 (46.7%) | 181 (56.7%) |  |
| Median Age (IQR) (years) | 8 (26.5) | 13 (25) | 15 (7) |
| No. Hispanic Ethnicity (%) | 4 (28.6) | 62 (20.2) | 987 (27.4) |
| No. Non-White Race (%) | 7 (46.7) | 53 (16.8) | 1015 (28.2) |
| **Individual Self-reported**<br>**Races** |  |  |  |
| American Indian or Alaska<br>Native (%) | 1 (6.7) | 0 (0.0) | 5 (0.14) |
| Asian (%) | 0 (0.0) | 9 (2.8) | 155 (4.3) |
| Black or African American<br>(%) | 0 (0.0) | 12 (3.8) | 662 (18.4) |
| White (%) | 8 (53.3) | 262 (82.1) | 2567 (71.3) |
| More than one race or | 6 (40.0) | 36 (11.3) | 193 (28.2) |
| Unknown (%) |  |  |  |

Over the course of the 12 months intervention period (see **Figure 1** for list of interventions), recruitment at our site increased from 15 subjects in the year pre-interventions to 319 in the following 12 months. The median age (IQR) at recruitment and percentage of female participants were 8 (26.5) years and 46.67%, respectively, between June 1, 2021 and May 31, 2022 and 13 (25) years and 56.74% between June 1, 2022 and May 31, 2023.

These descriptive statistics, along with the equivalent numbers for the overall clinic cohort from which subjects were recruited are shown in **Table 1**.

Total registrations by quarter were plotted over the two-year period, with different interventions noted in their corresponding quarter to show temporal relationship between interventions and recruitment rate (Figure 2). Notably, data about how participants learned about TrialNet (other than referring center) was not available.

**Figure 2.**
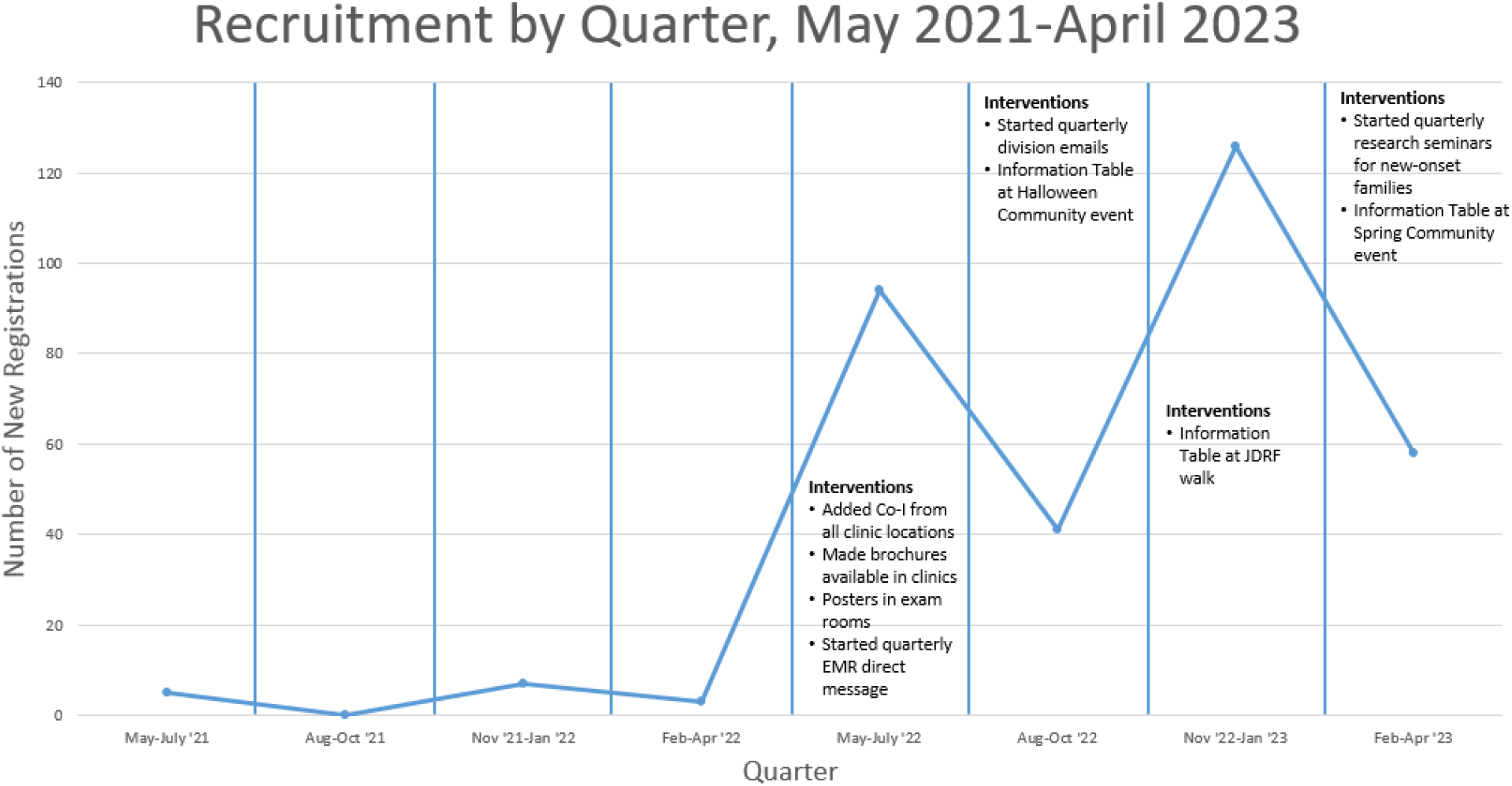
Participant enrollment over time, with dates of initial implementation of various recruitment strategies noted. Information tables at community events were events organized by our division for patients where a TrialNet investigator(s) and/or coordinator were present to advertise and assist with enrollment. Clinic messaging included posters and brochures made available to patients in exam rooms and/or at checkout. Division emails provided updates on recruitment and reminded faculty of available resources including EHR smart text and contact information for investigators and coordinator for additional help. EHR direct messages were sent to families of patients with T1D directly through the EHR patient portal with information about TrialNet and how to enroll.

## Discussion

The implementation of revised recruitment practices at our TrialNet site was associated with a notable surge in registrations over a 12-month period. These interventions were designed to address one common barrier to participation in research and clinical trials: lack of knowledge [7]. In our large center, with a large faculty group, only a small group are active investigators in TrialNet, with the rest of the faculty having differing levels of knowledge about the study, including how to refer interested individuals and answer questions about the study from those considering enrollment. To address this issue, our interventions aimed to 1) educate providers locally about TrialNet, its potential benefits to subjects, who and how to refer, and who to contact for more information or help; and 2) provide similar information directly to patients and their families from the investigator group. **Figure 1**, aside from listing the various interventions implemented at our center, groups them into which of these barriers they aimed to tackle.

While implementation of strategies to increase provider education and patient awareness was associated with an increase in recruitment rate overall and in each of the racial and ethnic group, the challenge of recruiting a representative study population remains. Based on the racial and ethnic breakdown of this site’s population of patients with T1D, the post-intervention study cohort does not completely mirror the affected population, namely non-White and Hispanic individuals. Previous studies such as that by Clark et al, have reported as barriers to enrollment in research such as lack of trust in the medical establishment and/or the biomedical research establishment affecting specific groups [7]. Our initial interventions implemented during this study period attempt to address the themes of lack of information, but are neither targeted to specific groups, nor do they address issues around fear or mistrust identified by Clark et al.

Specific interventions aimed at addressing some of these barriers to recruiting a diverse cohort are ongoing. Following the conclusion of the study period, for instance, we have implemented direct electronic health record (EHR) communication with information about TrialNet and contact information for study investigators and coordinator to the providers of patients from groups that remain underrepresented in our site’s enrollment efforts. In this manner, a provider who is known and trusted by potential participants and their families can introduce the study and directly provide information to the potential subject and their family.

Nationally, initiatives such as pre-set target enrollment rates for under-represented groups [8] have set the goal of recruiting a trial population that mirrors the affected population. The importance of such efforts in TrialNet is underscored by the approval of teplizumab, thus far the only disease-modifying therapy available for T1D [9]. Expanding recruitment and heterogeneity in studies like TrialNet can help to grow the scientific community’s knowledge of their pathophysiology, and to ensure such individuals are included in all stages of design, development, testing, and approval of therapies to slow or prevent onset of T1D.

While our study provides valuable insights into successful recruitment efforts, it is important to acknowledge limitations. Notably, we lack specific data attributing registrations to individual interventions due to the absence of information regarding how registrants first learned about TrialNet. Although we aligned interventions with registration trends over time, the precise influence of each intervention remains uncertain. Additionally, the reliance of self-reported data, including race and ethnicity, introduce potential biases. Despite these limitations, our study contributes significantly to understating of recruitment dynamics and underscores the need for continued refinement in recruitment practices. As mentioned above, rigorous statistical analysis was not performed due to the large differences in cohort sizes. In the future, if pace of recruitment is maintained, comparison of cohorts before and after interventions of specific strategies aimed at boosting recruitment of underrepresented groups may be possible.

In conclusion, our study documents a remarkable increase in TrialNet Pathway to Prevention Study participation subsequent to implementation of various initiatives. Despite persistent disparities in proportional enrollment rates among certain racial/ethnic groups, there was a substantial increase in the number of participants across nearly all demographic categories. The strategies presented in this study hold promise for enhancing recruitment numbers and diversity in other centers. However, further research is warranted to elucidate underlying factors contributing to the disproportionate research participation and addressing them among different racial/ethnic groups.

## Data Availability

All data produced in the present study are available upon reasonable request to the authors, with consent as needed, by TrialNet.

